# Relative risks, the probability of necessity, and attributable fractions

**DOI:** 10.1101/2024.07.03.24309898

**Authors:** Anthony J. Webster

## Abstract

Epidemiologists are careful to describe their findings as “associations”, and to avoid any causal language or claims. Arguably, this attempt to avoid reference to causal processes has become counterproductive. Explicitly stated or not, assumptions about causal processes are inherent in the formulation and interpretation of any statistical study. This article offers a bridge between established, extensively developed proportional hazard methods that are used to study longitudinal observational cohort data, and results for causal inference. In particular, it considers the burden of disease that would not have occurred, but for an exposure such as smoking. It shows how this “probability of necessity”, relates to population attributable fractions, and how these quantities along with their confidence intervals, can be estimated using conventional proportional hazard estimates. The example may often apply to cohort studies that consider disease-risk in the absence of prior disease. More generally, equivalent estimates can often be constructed when there is sufficient understanding to postulate a model for the causal relationship between exposures, confounders, and disease-risk, as summarised in a directed acyclic graph (DAG).

## 1 Causal assumptions are necessary for designing and interpreting an analysis

Concerns about reporting epidemiological data without clearly stated causal assumptions have been raised before [1, 2], with authors emphasising that results risk being misinterpreted. This has been referred to as the “Table 2” fallacy [1], because statistical associations are typically reported in table 2 of most epidemiological articles. Associations are often reported without a clear distinction between estimates that are intended to describe causal associations, and those that are used for adjustment, whose causal interpretations are often unclear. A clear statement of causal assumptions has several important advantages. Most obviously it helps the reader, who can assess and interpret studies from the perspective of the author. But there is another benefit, that a causal diagram allows a far broader range of inferences to be made using new results for causal inference [3, 4].

In contrast to observational epidemiology, it is standard practice for Mendelian Randomisation studies to report the assumed causal structure as a directed acyclic graph (DAG) [5]. It is helpful to compare the two approaches. For observational studies, epidemiologists delve into the literature and try to ascertain a picture of risk-modifying factors and the relationships between them. This informs their statistical analysis and its interpretation, but much of that is done implicitly without stating any assumed causal relationships needed for causal interpretation of the results. In Mendelian Randomisation studies (MR), epidemiologists search for biological reasons to select genetic variants that are likely to modify disease risk solely through the exposure that they are interested in (“instrumental variables” [5]). However, in MR studies epidemiologists state their assumed causal diagram for the relationships between genetic variants, exposures, confounders, and disease. This makes the assumptions clear, making it easier to identify when they fail, and helping to clarify how results should be interpreted.

## 2 Causal assumptions, allow causal inferences

The process of formulating a causal hypothesis such as a DAG, is familiar to a theoretical physicist - you state the assumptions in your physical model and then make your deductions. If the assumptions need modifying then the model is changed, and your deductions are updated. Whether stated explicitly or not, observational epidemiology relies on causal relationships to plan and interpret statistical analyses. Once these working hypotheses are stated then deductions can be made using mathematical methods, in a similar way to Mendelian Randomisation or theoretical physics. A DAG then allows methods for causal inference to tackle questions that would otherwise be inaccessible. For example, you can ask the following questions regarding risks associated with smoking [3, 4]:

1. If everyone in the population stops smoking, how much will disease incidence change? This question is associated with population health, and involves measures such as population attributable fraction (PAF), excess fraction, and average causal effect (ACE).
2. If you stopped smoking, how much will your disease risk change? This question is related to your individual health, and involves measures such as the effect of treatment on the treated (ETT).
3. Given that you died from lung cancer, how likely would this have been if you had not smoked? This question is familiar in legal contexts, where it translates as the probability an event would not occur, but for the defendant’s actions, and is often referred to as the probability of necessity (PN).

Although these questions can often be precisely defined, and their analysis usually leads to distinctly different quantities, there are relationships between them [3, 4]. In particular, this article considers the relationship between probability of necessity (PN), and a recently developed population attributable fraction (*A*_*f*_) that was intended to account for confounding with measured confounding factors [6]. It is shown that when studying exposures that can solely increase disease risk, an assumption known as “monotonicity” [3], then PN is proportional to *A*_*f*_. Furthermore, it was previously shown how *A*_*f*_ can be evaluated for a conventional longitudinal study with a proportional hazard estimate of relative risks, making *A*_*f*_ comparatively easy to evaluate. The next section considers PN, attributable fractions, and the relationship between them.

## 3 Probability of necessity and attributable fractions

The probability of necessity is intended to estimate the probability that a disease would not have occurred, but for the exposure (such as smoking), having occurred. For completeness, the formal definition in terms of counterfactual notation is given below,

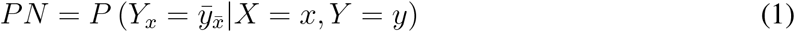

where *X* is a binary risk factor such as smoking status, that can take values *X* = 1 or *X* = 0, *Y* is the disease status that with *y* true corresponds to disease having occurred by age *t* and 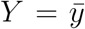 (false) if it has not. The bar in e.g. 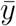 is used to indicate its negation from true to false, or “not” *y*, and the subscripted *Y*_*x*_ indicates a “counterfactual” scenario where *X* is taking the specific value *X* = *x*. The interpretation and analysis of these types of expressions takes some practice, but here we can quote and use established results. In particular, if we consider diseases where an exposure such as smoking can only increase risk (an assumption that is referred to as “monotonicity”), then PN becomes [3, 4],

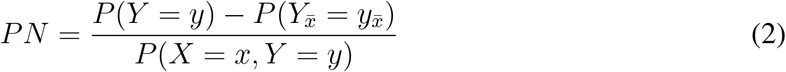

where 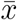 corresponds to *X* = 0 (false), and *x* to *X* = 1 (true). This may be written as,

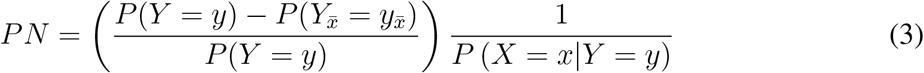

The first term on the right-side is identical to the population attributable fraction *A*_*f*_ defined and studied by Webster [6]. This makes sense. The attributable fraction *A*_*f*_ was intended to describe the proportion of disease that could be avoided in a population if the exposure did not occur, and is important for prioritising public health initiatives. Rearranging terms in Eq. 3 we can write,

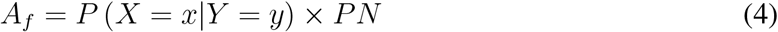

meaning that *A*_*f*_ will be small unless both PN and the proportion of people with the disease who have been exposed *P* (*X* = *x*|*Y* = *y*), are both reasonably large. Arguably, from a public health perspective, it is more important to identify exposures that have the greatest overall impact on the population.

Despite the potential usefulness of PN and *A*_*f*_ for characterising the avoidable diseases that are caused by exposures such as smoking, they are rarely reported by analyses of observational cohort studies. This may be because they are not yet widely known and understood, but is also because there are no established methods to calculate them. However for a simple but widely assumed DAG (figure 1), it was recently shown how *A*_*f*_ can be estimated using conventional proportional hazards analyses, leading to,

**Figure 1:**
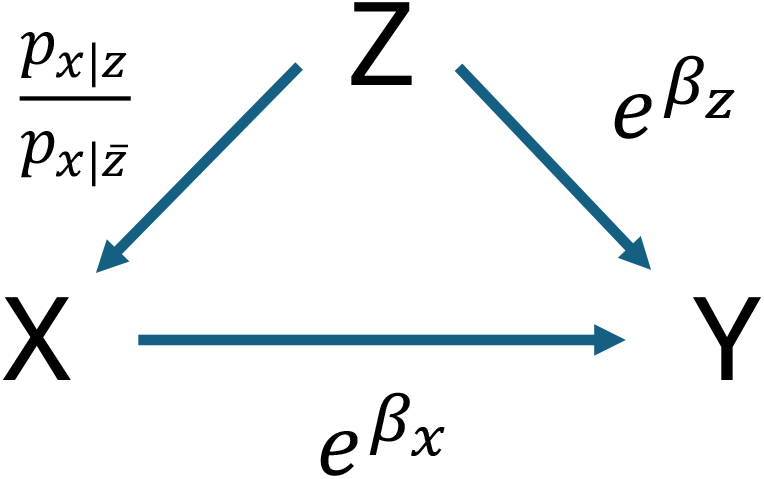
A simple DAG is considered, that may often be a reasonable or implicit assumption for an observational study. Relative risks are indicated by 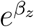 and 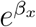 respectively, and we also specify the relative risk of exposure *X* = 1 given *Z* = 1, compared with *Z* = 0. Simulated data are created using *p*_*x*_, *p*_*z*_, and the relative risks 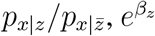, and 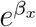, (as detailed in Appendices A and B).

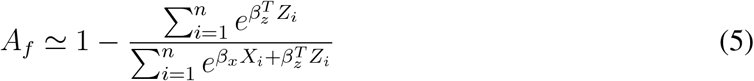

where 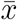 was denoted by *X* = 0. Eq. 5 allows *PN* to be estimated from *A*_*f*_, provided that *P* (*X* = *x*|*Y* = *y*) can be adequately estimated from the proportion of people with the disease who have been exposed (e.g. to smoking). In Eq. 5, *β*_*z*_ is a vector of parameters to adjust for all risk and confounding factors, other than *X*, and the sum over *i* includes all *n* individuals in the cohort being studied. Note that correlations between the risk factors *X*_*i*_ and other parameters *Z*_*i*_, are captured in the denominator of Eq. 5. The analysis assumes:

1. Causal model: Eq. 5 was calculated for the simple causal diagram in figure 1 with exposures *X*, and confounders *Z* that modify the probability of both *X* and disease *Y*.

More generally, given a causal diagram, it is expected that established methods from the causal inference literature [3, 4] can be combined with the approximations outlined here to calculate analogous expressions.

2. Low disease risk: For most diseases in most people without pre-existing disease, disease risk is very small for an average UK lifespan of approximately 81 years [6, 7].

This surprisingly little-known fact about disease risk [6, 7], allows cumulative density functions and probability density functions to be approximated by the cumulative hazard [8] and hazard function [8] respectively. This greatly simplifies the analysis by removing the age dependence from the attributable fraction *A*_*f*_ (and consequently also for PN). Alternately, *A*_*f*_ can be regarded as exact, but defined in the theoretical limit of small enough age [6].

3. Statistical model: The proportional hazards analysis must model the data sufficiently well, and adjust for all relevant parameters.
4. There is a sufficiently large cohort size *n*, with sufficient numbers in each category to allow integrals to be approximated by sums over individuals in the data [6].

The assumptions above are explored further later. In addition, Eq. 2 is limited to:

5. Binary exposures that increase risk (“monotonicity”).

For example, if assessing the influence of smoking on COPD, we would assume that if a non-smoker developed COPD, then they would have also developed COPD if they had smoked. With greater expertise a more general range of questions can be tackled, and results can be used that allow some assumptions to be relaxed. However the examples above already allow estimates for a substantial number of important diseases and exposures such as smoking [6].

## 4 Confidence intervals and estimate accuracy

The approximation of integrals with sums is justified by the law of large numbers [9], and will be reasonable for large cohorts such as UK Biobank [10], provided categorical variables have reasonably balanced proportions of people in each category. Because of the central limit theorem, Eq. 5 for *A*_*f*_, involves a ratio of normally distributed variables whose variances tend to zero as *n* → ∞. Because the means of the numerator and denominator are non-zero, and their variances tend to zero with *n* → ∞, then as explained more fully in Appendix C, Eq. 5 will have a normal distribution for sufficiently large *n*. This and the finite variance of the estimate allow the approximation’s accuracy to be assessed with bootstrap sampling [9]. The procedure for a statistic *S*({*X*_*i*_, *Z*_*i*_}) of the data, is to sample with replacement *n* individuals from a dataset of size *n*, calculate the sum *S*({*X*_*i*_, *Z*_*i*_}), repeat this *B* times and estimate the variance from, *v*_*boot*_ = (1*/B*) Σ_*j*_ (*S*_*j*_ − (1*/B*) Σ_*k*_ *S*_*k*_)^2^, where *S*_*j*_ is the value of the sum using the *j*th sample [9]. For the examples in this article, *B* = 500.

Appendices A and B show how realistic cohort data can be simulated for a DAG corresponding to figure 1, and how the attributable fractions can be exactly calculated. This allows the method to be tested. Figure 2 compares the estimated and exact *A*_*f*_, and *PN*, versus the median age in the cohort. The method is expected to fail for high median ages, but remains good for ages ∼ 80 years, which is similar to the average UK life expectancy. Therefore the estimates are expected to be reasonable for typical UK cohorts. Further simulations with 1 and 2 million individuals (Appendix E), found very similar results.

**Figure 2:**
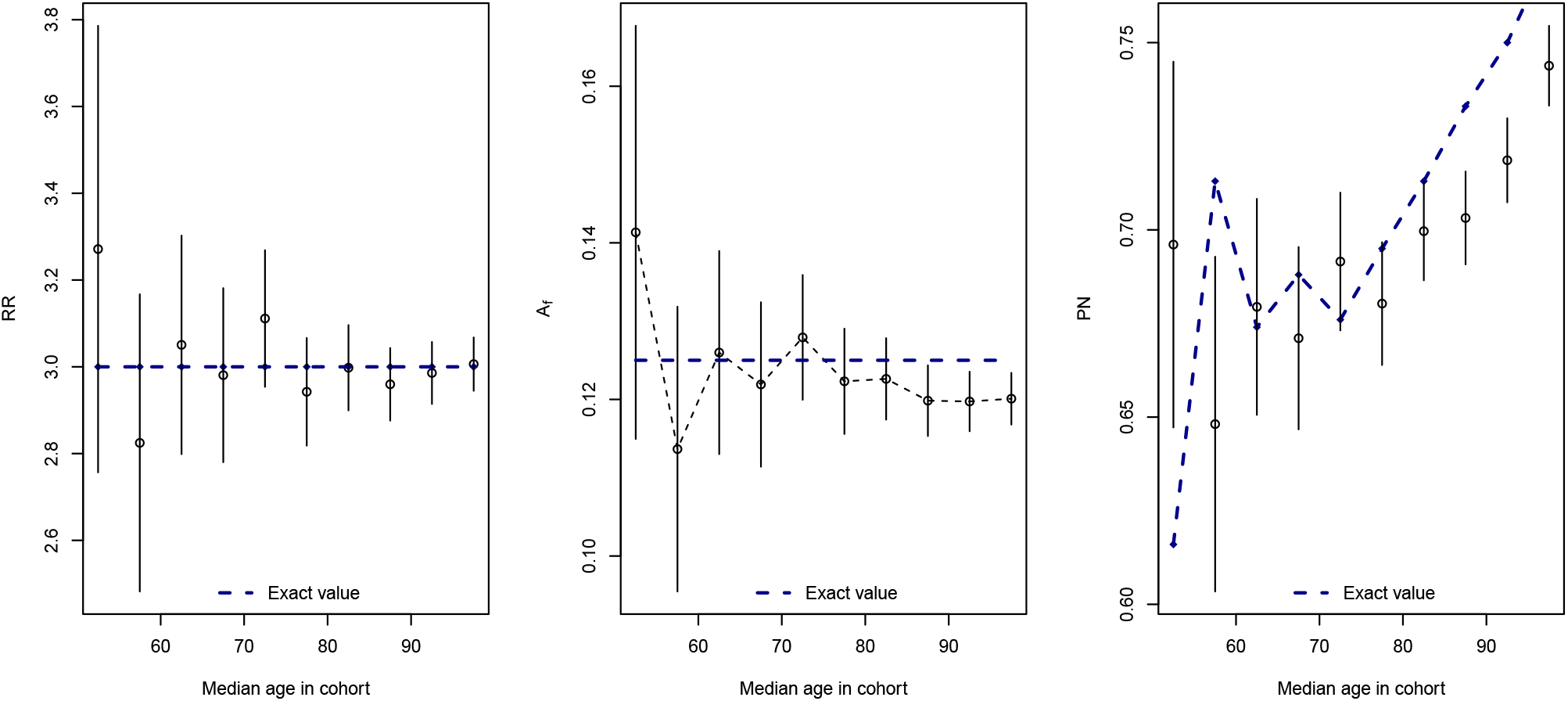
*A*_*f*_ can be exactly calculated for simulated data (Appendices A and B), allowing *P* (*X* = *x*|*Y* = 1) and the probability of necessity PN to be estimated using Eq. 4 and the data. As the cohort’s median age increases, Eq. 5’s approximation for *A*_*f*_ starts to fail. Reassuringly however, the exact value remains within the 95% confidence intervals for ages greater than the median UK life expectancy (79 and 83 years for men and women in born in 2018-2020).

Figures 3 and 4 compare estimated and exact *A*_*f*_ and *PN* for a simulated range of data generated by a causal process described by figure 1. The results are also listed in Table 1. Details of how the data were generated are in appendices A and B, and involve a similar type of cohort to UK Biobank, with a similar number of ∼ 500, 000 individuals. The examples start with “No effects” with relative risks of 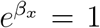 and 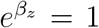, with “Exposure” or “confounding” corresponding to 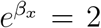 or 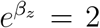 respectively, and “Strong exposure” or “strong confounding” corresponding to 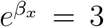 or 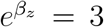 respectively. Exact and estimated attributable fractions *A*_*f*_ and *PN* are compared in figures 3 and 4, for all combinations of 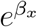 and 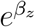 in {1, 2, and 3}. The exact *A*_*f*_ is comfortably within the 95% confidence intervals, and although there is slightly more variation of estimates for *PN*, all but “Exposure only” have values within the confidence intervals.

**Table 1:** For simulated data, the table lists estimates for different types of attributable fractions. RR_0_ and A_*f*0_ are calculated exactly using parameters that specify the simulated data, and PN_0_ is calculated using an estimate for *P* (*X* = 1|*Z* = 1). “Exposure”, “Strong exposure”, have 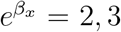, 3, and “Confounding”, “Strong confounding”, have 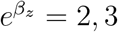.

|  | RR<sub>0</sub> | RR | A<sub>f0</sub> | Af | PN<sub>0</sub> | PN | ERR | A<sub>RR</sub> |
| --- | --- | --- | --- | --- | --- | --- | --- | --- |
| No effects | 1.00 | 0.97 [0.88,1.07] | 0.00 | 0 [0,0.01] | 0.00 | 0 [0,0.1] | 0 [0,0.09] | 0 [0,0.1] |
| Confounding only | 1.00 | 1.01 [0.94,1.08] | 0.00 | 0 [0,0.01] | 0.00 | 0.01 [0,0.08] | 0.22 [0.16,0.27] | 0.01 [0,0.08] |
| Strong confounding only | 1.00 | 1.04 [0.97,1.1] | 0.00 | 0.01 [0,0.01] | 0.00 | 0.04 [0,0.1] | 0.32 [0.28,0.36] | 0.04 [0,0.1] |
| Exposure only | 2.00 | 2.11 [1.97,2.26] | 0.09 | 0.1 [0.09,0.11] | 0.49 | 0.53 [0.5,0.56] | 0.52 [0.49,0.55] | 0.53 [0.49,0.56] |
| Exposure and confounding | 2.00 | 2.03 [1.91,2.14] | 0.11 | 0.11 [0.1,0.12] | 0.51 | 0.52 [0.49,0.54] | 0.61 [0.59,0.63] | 0.51 [0.48,0.53] |
| Exposure and strong confounding | 2.00 | 1.97 [1.88,2.06] | 0.12 | 0.12 [0.11,0.13] | 0.52 | 0.5 [0.48,0.53] | 0.64 [0.62,0.66] | 0.49 [0.47,0.52] |
| Strong exposure only | 3.00 | 3.09 [2.89,3.29] | 0.17 | 0.17 [0.16,0.18] | 0.67 | 0.69 [0.66,0.71] | 0.67 [0.65,0.69] | 0.68 [0.66,0.7] |
| Strong exposure, and confounding | 3.00 | 2.94 [2.8,3.07] | 0.20 | 0.19 [0.18,0.2] | 0.70 | 0.68 [0.66,0.7] | 0.73 [0.72,0.74] | 0.66 [0.64,0.68] |
| Strong exposure and strong confounding | 3.00 | 3.06 [2.93,3.18] | 0.21 | 0.22 [0.21,0.23] | 0.68 | 0.7 [0.68,0.71] | 0.77 [0.76,0.78] | 0.67 [0.66,0.69] |

**Figure 3:**
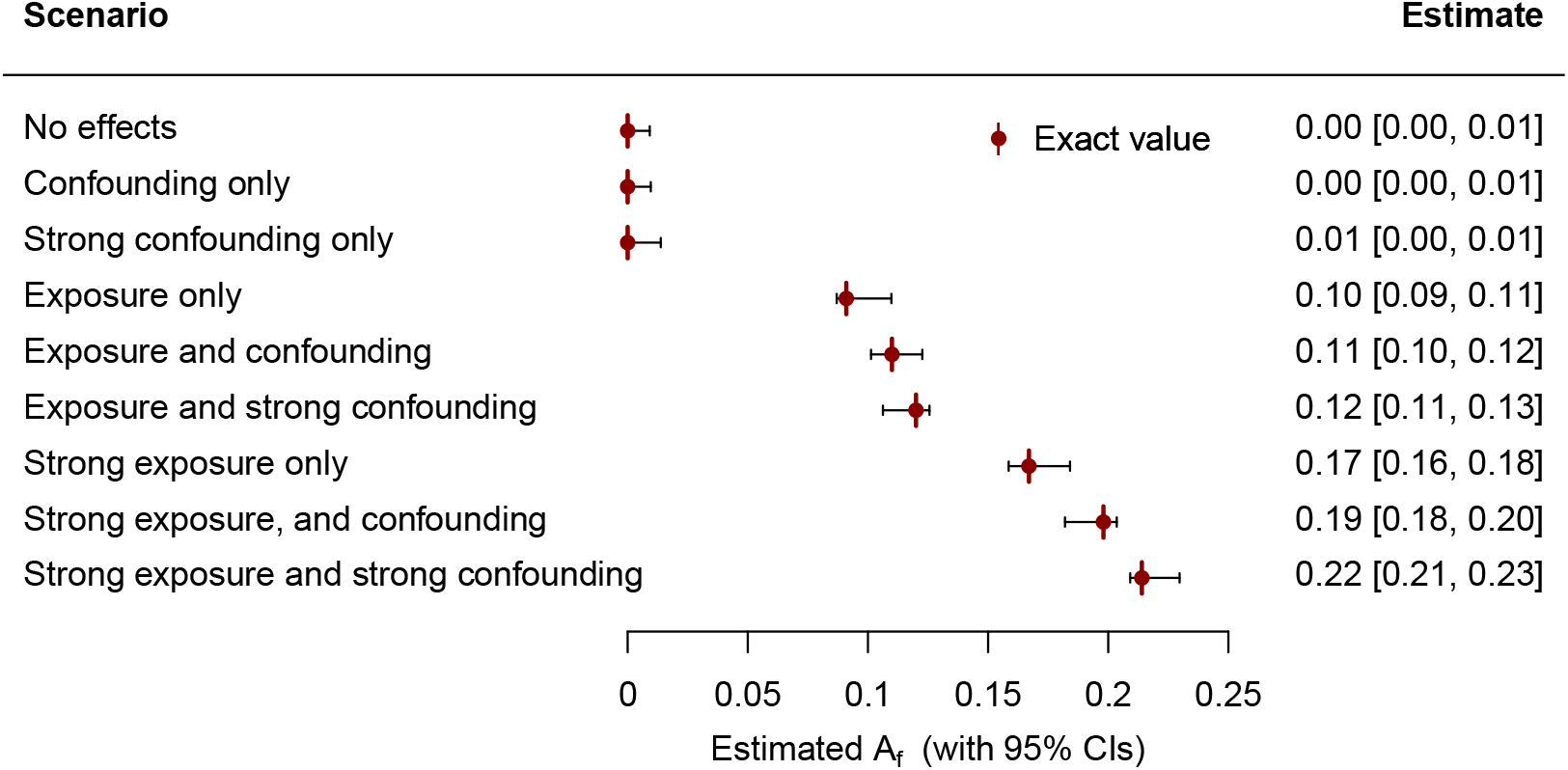
Simulated data for ≃ 500,000 individuals were created for scenarios with relative risks of 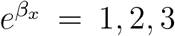 (“no exposure” to “strong exposure”), and 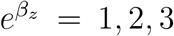 (“no confounding” to “strong confounding”). Eq. 5 was used to estimate *A*_*f*_, with confidence intervals estimated using the bootstrap (see text for details).

**Figure 4:**
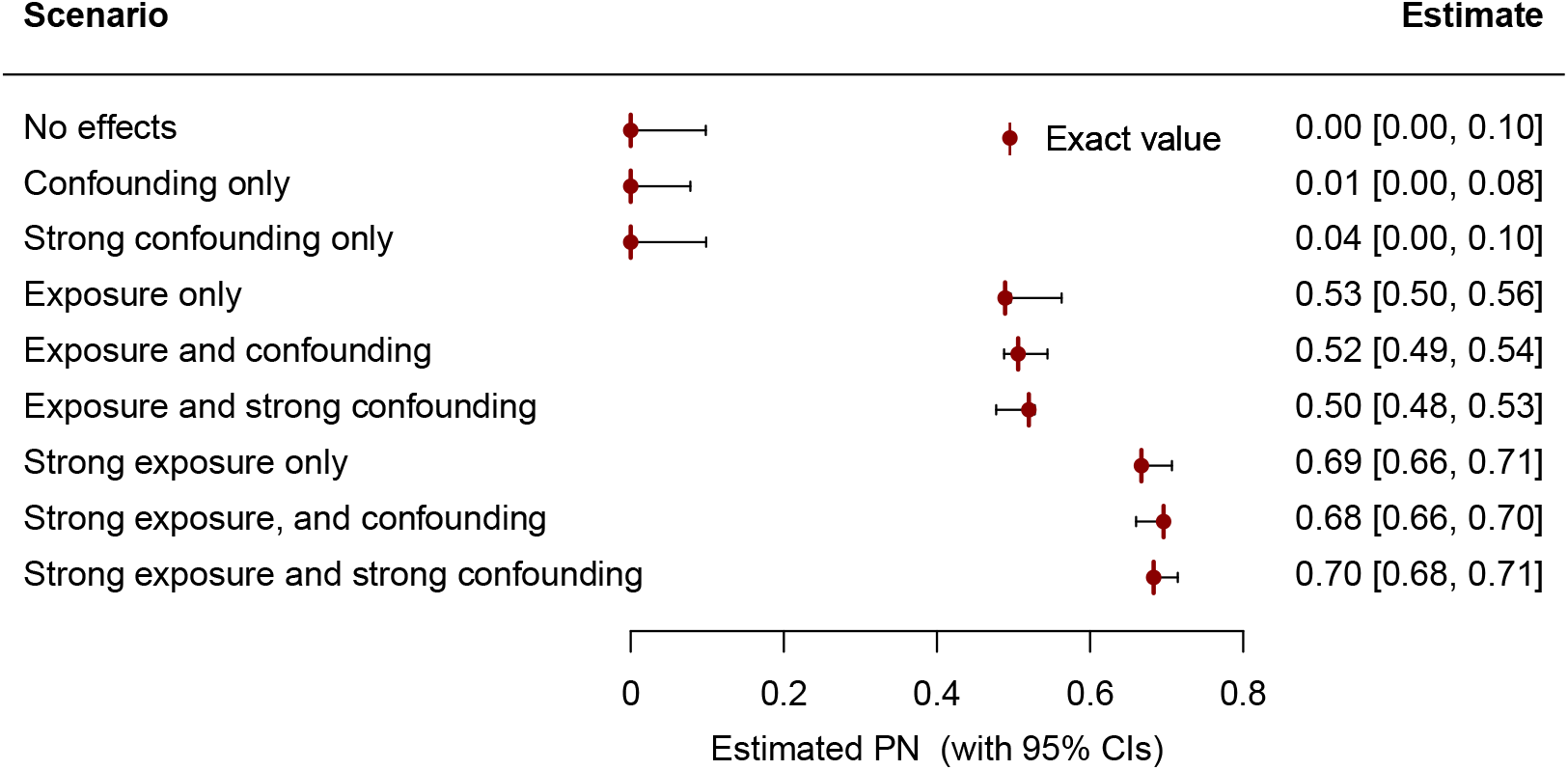
Simulated data for ≃ 500,000 individuals were created for scenarios with relative risks of 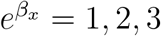 (“no exposure” to “strong exposure”), and 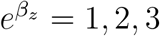 (“no confounding” to “strong confounding”). Eq. 5 was used to estimate *A*_*f*_ and (subsequently) *PN*, with confidence intervals estimated using the bootstrap (see text for details).

## 5 Other attributable fractions

At least two other attributable fractions are often discussed, and are briefly considered here. The excess risk ratio (ERR) [3, 4], is defined as,

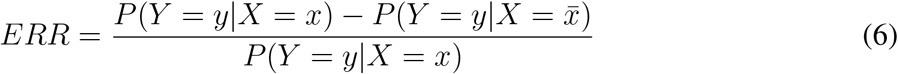

Appendix D shows how this can be estimated in a similar way to *A*_*f*_, leading to,

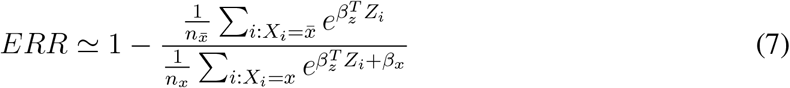

where as for Eq. 5, *x* is denoted by 1 and 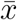 is denoted by 0, so that 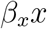 equals *β*_*x*_ and 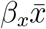 equals 0. There are important differences between ERR and *A*_*f*_. For ERR the sums are over subsets of the population that have *X*_*i*_ = *x* and 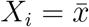 respectively, and in the denominator for ERR *X*_*i*_ = 1 whereas in the denominator of *A*_*f*_ there is a sum over all *X*_*i*_ (that can have both *X*_*i*_ = 0 and *X*_*i*_ = 1). Similarly to the approximations for *A*_*f*_, Eq. 7 is approximately equal to Eq. 6 for most of the lifetime of most people in the UK, or can alternately be regarded as a definition of ERR for a sufficiently young and healthy population with no pre-existing diseases (that could modify risk of the disease being studied).

Another commonly seen attributable fraction for a risk factor *x* with relative risk 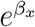, is,

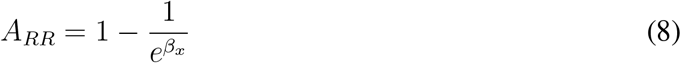

*A*_*RR*_ is proportional to the difference in probability of an individual getting a disease when exposed, compared to not exposed, when subject to the same confounding factors [6]. This factor is analogous to the effect of treatment on the treated (ETT) [3, 4, 6], but now “treatment” is an exposure that increases risk, and we measure how much the risk is increased (as opposed to decreased). This is more relevant for individual risk, but its interpretation as an attributable fraction is unclear when dealing with an individual, as opposed to a population.

Although widely discussed, the greatest limitation of ERR is that it does not have a causal interpretation (except in terms of other quantities [3, 4]). For the examples listed in Table 1, ERR has values that are very different to the other causally interpretable quantities, so it does not in general provide a reliable estimate for any of those. As discussed elsewhere [6], and described above, A_*RR*_ has a causal interpretation that is related to ETT and an individual’s increased disease risk. In the examples considered here, A_*RR*_ had similar values to PN.

## 6 Which attributable fraction should you report?

Whether to report an attributable fraction, a relative risk, a probability of necessity, or some other causally meaningful quantity, depends on what you are intending to quantify. It might be that several of these might be reported together, because they each characterise different aspects of an exposure’s impact on disease risk. To help with this, several quantities are summarised below:

### RR

The estimated hazard of disease in an exposed individual, relative to someone that is not exposed. This is approximately equal to the ratio of probability for disease in an exposed, compared with the unexposed [6, 7].

### A_f_

The proportion of disease that occurs in the population being studied (typically a cohort), that could have been prevented if all the population had avoided the exposure. This is a factor *P* (*X* = 1|*Y* = 1) smaller than PN, because disease incidence can only be reduced for the proportion of the population that are exposed.

### PN

The probability that a disease would not have occurred, but for the exposure [3, 4]. For the examples in table 1, this increased with greater relative risks of the exposure or confounding factors associated with increased exposure (see figure 4 and table 1).

### ERR

Does not have a causal interpretation, except in terms of other quantities [3, 4]. For the examples in table 1, its values usually differed from the other causally interpretable quantities, so it would not in general provide an approximation for them.

### A_RR_

The probability of an individual getting a disease when exposed, relative to someone without the exposure, but with the same confounding factors. It has a causal interpretation that is analogous to effect of treatment on the treated [6], but its interpretation as an attributable fraction is unclear because it refers to an individual, not a population. For the examples in table 1, it shows a similar trend to PN, but with smaller estimates when confounding is present.

## 7 Limitations of directed acyclic graphs (DAGs)

Despite the many advocates for a greater use of DAGs in epidemiology [1–4, 12], several authors have cautioned against over-reliance on them, highlighting important examples where their value is limited [13, 14]. For example, what DAG should you formulate when considering the influence of BMI on ill health? It may differ depending on whether you are interested in understanding the biologically-mediated causes of ill health, or the underlying societal issues that lead to higher BMI [14]. This and other examples highlight the limitations of formal causal reasoning, and suggest that causal methods should be regarded as tools to provide insights, similar to how a physicist explores the physical world using mathematical models.

An important purpose of this article is to highlight opportunities that arise from causal assumptions that are often made in the course of many epidemiological studies. The notion of using the best available information to form a model, and then exploring its predictions, is a legitimate and established mode of study in the physical sciences. It is similar to “inference to the best explanation” [14], where a DAG may represent our best understanding, and subsequently allows a more nuanced interrogation of the data. DAGs and causal inference are valuable tools, but causal understanding emerges from a body of evidence arising from several distinct sources [15, 16]. The approach is familiar to physicists, where a theoretical model is formulated, its consequences explored, and the model revised when new data make it necessary to do so.

## 8 Summary and conclusions

It has become standard practice for observational studies to report relative risks [11], most commonly calculated with proportional hazards methods. There is considerable experience with these widely used methods, that are thought to produce reliable results that can be reproduced in different cohorts. However the reporting of studies using them (and other methods), have been criticised for failing to identify which parameters are risk factors or confounders, or the causal relationships that have been assumed to hold between them [1, 2]. This is unlike Mendelian Randomisation, in which a DAG is used to indicate causal relationships. However, whether causal modelling assumptions are explicit or not, they are necessary for formulating and interpreting the results of a statistical analysis [1]. When stated explicitly in the form of a DAG, a causal diagram clarifies assumptions for the reader, but also allows a broader range of methods from causal inference to be used, similar to how a theoretical physicist will state a model and then make deductions. This allows a much wider range of causally meaningful quantities to be calculated, such as population attributable fractions and the probability of necessity. These offer valuable alternative characterisations of an exposure’s influence on a population or individual, that ideally would be regularly reported.

The approach is illustrated for scenarios with the DAG in figure 1, for which it is shown how attributable fractions and the probability of necessity can be estimated using conventional proportional hazards methods. This allows decades of experience that epidemiologists have gained with using proportional hazards, to be used for the accurate calculation of these alternative measures of risk. The estimates were found to be reliable for the wide range of simulated data that were considered, that were designed to be similar to that expected for common diseases in cohorts such as UK Biobank. More generally, for studies where a DAG can be formulated with reasonable confidence, it is expected that the approach can be applied using methods from causal inference [3, 4], leading to similar but modified versions of Eq. 5. In future, epidemiologists will become more familiar with causal quantities such as PN, and with using methods from causal inference to formulate estimates. This will allow a wider range of questions to be tackled and provide alternative measures of risk, improving our understanding of the causes of diseases and the consequences of interventions.

## Data Availability

Simulated data are used in the manuscript. Code to produce data and figures will be made available to referees and following publication.

## Acknowledgements

No funding was received for this work, that was completed while employed at Oxford University. Thanks to the causal inference groups at the Turing Institute and Oxford University’s Statistics Department for feedback and encouragement, in particular to their seminar organisers Peter Tennant, Max Little, Robin Evans, Frank Windmeijer, Xi Lin, and Vik Shirvaikar.

## A Example - Exact values for *PN* and *A*_*f*_

Using 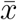 to denote the negation of *x*, Eqs. 3 and 4 give,

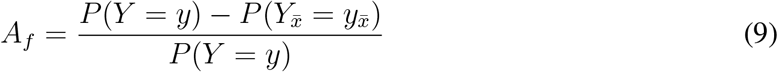

That for the example DAG of figure 1, the adjustment formula [3, 4] can be used to write it as,

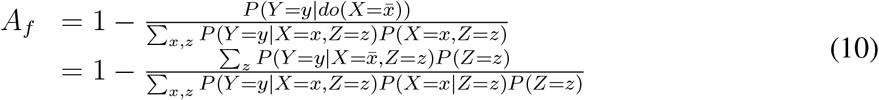

To simplify the notation, let,

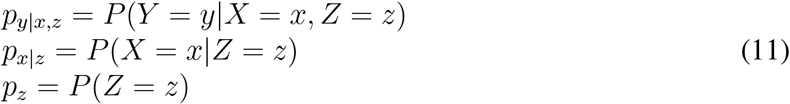

with equivalent expressions for negations of *x*, so that if *x* replaces 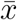 then *p*_*x*_ is replaced by 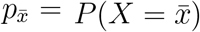. Then,

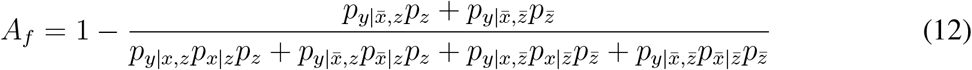

Noting that 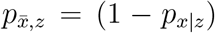 and 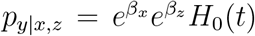 where *H*_0_(*t*) is the cumulative baseline hazard function, and similarly for the other terms, then,

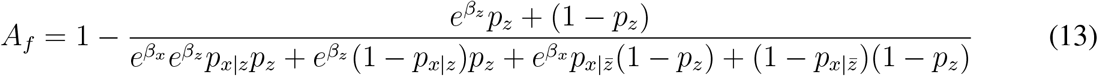

where the factors of *H*_0_(*t*) have cancelled in the fraction. Therefore, specifying the relative risks 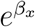 and 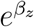, and 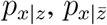, *p*_*x*_, *p*_*z*_, determines *A*_*f*_. Note that if 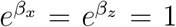, then *A*_*f*_ = 0, as it should do. Hence we can simulate data using 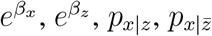, *p*_*x*_, and *p*_*z*_, and test the estimate of Eq. 5 by comparing it with the exact value given by 13.

In practice we only need to specify 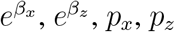, and the relative risk 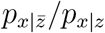. To see why, note that,

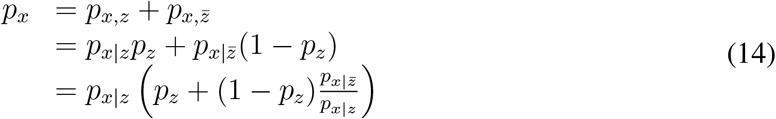

giving,

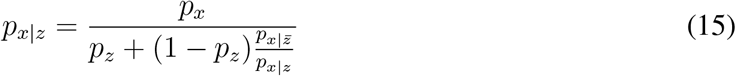

Similarly, we can rearrange,

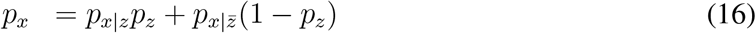

to give,

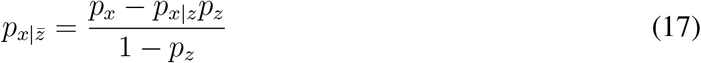

with *p*_*x*|*z*_ evaluated using Eq. 15.

## B Simulated data

Consider the simplified DAG of figure 1, with exposures *X* and confounders *Z*. For this example we will consider *X* as smoking status of yes or no, and *Z* as whether you live in a city or the country. The example is intended to test the statistical approach, and not necessarily to represent a real situation. Data are simulated by:

1. Specify the size *n* of the simulated dataset.
2. Specify *p*_*z*_ and the desired *p*_*x*_, and simulate whether each individual lives in the city (*z*) or country 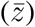.
3. Specify the relative risk 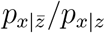 for smoking status if you are not in a city, compared with if you are.
4. Using *p*_*x*|*z*_ and 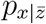 from Eqs. 15 and 17, simulate whether individuals smoke, given their city residence status.
5. Simulate the age of joining the cohort, here taking a minimum age of 45 plus a random number of years in (0, 20).
6. Simulate their age at the end of the study period, as their age at the study start plus 10, plus a random number of years in (0, 15).
7. Using smoking and membership status as risk factors with specified relative risks 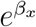 and 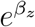, simulate an age of disease onset using a Weibul model with scale factor 130 and shape factor 8 (chosen to give similar disease incidence to a common disease).
8. If the age of disease is before the study period starts or within 1-year of the study start, exclude the data to simulate the process of trying to reduce the (hypothetical) risk that a person joined the study due to having disease. If the disease onset is after the end of the study period then censor.

This now provides a data set with ages *t*_*start*_, *t*_*end*_, and status (censor or not), plus smoking status and city membership. The simulated data can then be fit with a proportional hazards model (the Weibull distribution is a specific type of proportional hazards model), and the estimates compared with the known relative risks and attributable fraction given by Eq. 13. Table 1 provides comparisons between the estimated and actual relative risks and attributable fractions. In the examples,

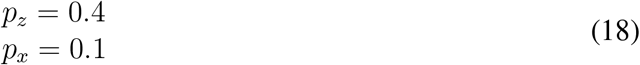

so that the probability of an exposure will be about 10%, and the probability of *Z* = 1 is slightly less than *Z* = 0 (approximately 40%). The probability of *X* = 1 if *Z* = 0 was taken to be a factor 0.25 smaller than the probability of *X* = 1 if *Z* = 1, with,

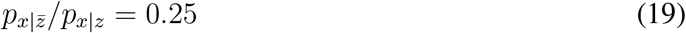

giving the probability of an exposure approximately 4 times greater if *Z* = 1. The relative risks took values of,

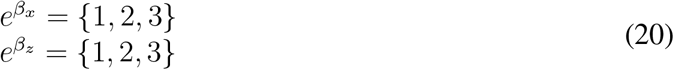

## C *A*_*f*_ has a normal distribution

Eq. 5 involves the ratio,

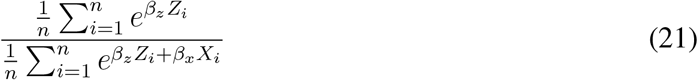

The expectation and variance of the numerator are 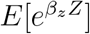 and 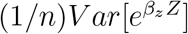 respectively, and for the denominator 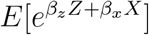 and 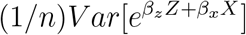. By the central limit theorem, the numerator and denominator are normally distributed, with expectation and variance as given. If the means were zero then the ratio would have a Cauchy distribution, however because the variances tend to zero as *n* → ∞ and their means are positive and non-zero, then the ratio will tend to a normal distribution [17], with,

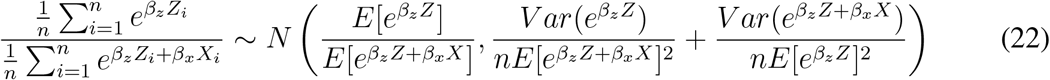

## D The excess risk ratio (ERR)

Another form of attributable fraction is the excess risk ratio (ERR) [3, 4], defined as,

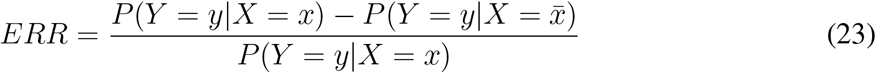

This can be estimated in a similar way to *A*_*f*_.

Recall that *y* true implicitly corresponds to the statement that disease has occurred by an age less than *t*, and *y* false is that disease has not yet occurred by age *t*. Then as in the derivation of Eq. 5 for *A*_*f*_ [6], *P* (*Y* = *Y* |*Z* = *z, X* = *x*) = *F* (*t*; *z, x*), where *F* (*t*; *z, x*) is the cumulative distribution function for disease by age *t*, and we make the assumption 2, that this can be approximated with a proportional hazards model with 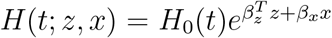. Then to approximate Eq. 23 with Eq. 26, we write,

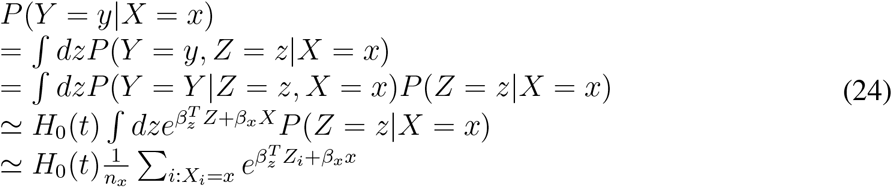

where the 3rd line takes 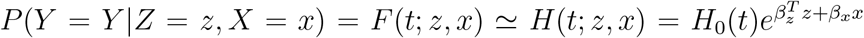, and as before [6] it is assumed there are sufficient data to be able to approximate the integral with a sum over the data in the population (assumption 4). Note that the last line has denoted *x* true as *x* = 1, and that the sum is over the subset of individuals for which *x* is true. The number of data with *X* = *x*, and 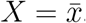, are denoted by *n*_*x*_ and 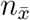 respectively. Similarly,

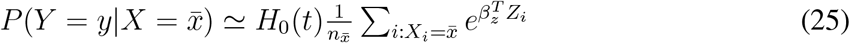

where now *x* is false is denoted as *x* = 0, and the sum is over all individuals for which *x* is false. Using Eq. 6 along with Eqs 24 and 25, gives,

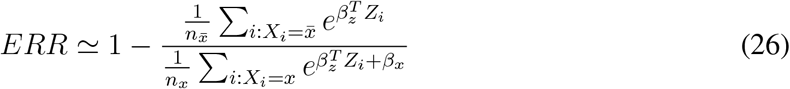

where as for Eq. 5, *x* is denoted by 1 and 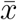 is denoted by 0, so that *β*_*x*_*x* equals *β*_*x*_ and 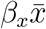 equals 0. There are important differences between ERR and *A*_*f*_. For ERR the sums are over subsets of the population that have *X*_*i*_ = *x* and 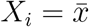 respectively, and in the denominator for ERR *X*_*i*_ = 1 whereas in the denominator of *A*_*f*_ there is a sum over all *X*_*i*_ (that can have both *X*_*i*_ = 0 and *X*_*i*_ = 1). Similarly to the approximations for *A*_*f*_, Eq. 7 is approximately equal to Eq. 6 for most of the lifetime of most people in the UK, or can alternately be regarded as a definition of ERR for a sufficiently young and healthy person with no pre-existing disease.

## E Additional examples

Figure 5 shows similar tests to those in in 2, but now with samples of 1 million and 2 million individuals respectively, as opposed to 500,000 individuals in figure 2. As for figure 2 the results are reassuringly accurate for cohorts with a median age that is likely to be seen in practice, in addition, they appear to become more accurate with increasing sample size.

**Figure 5:**
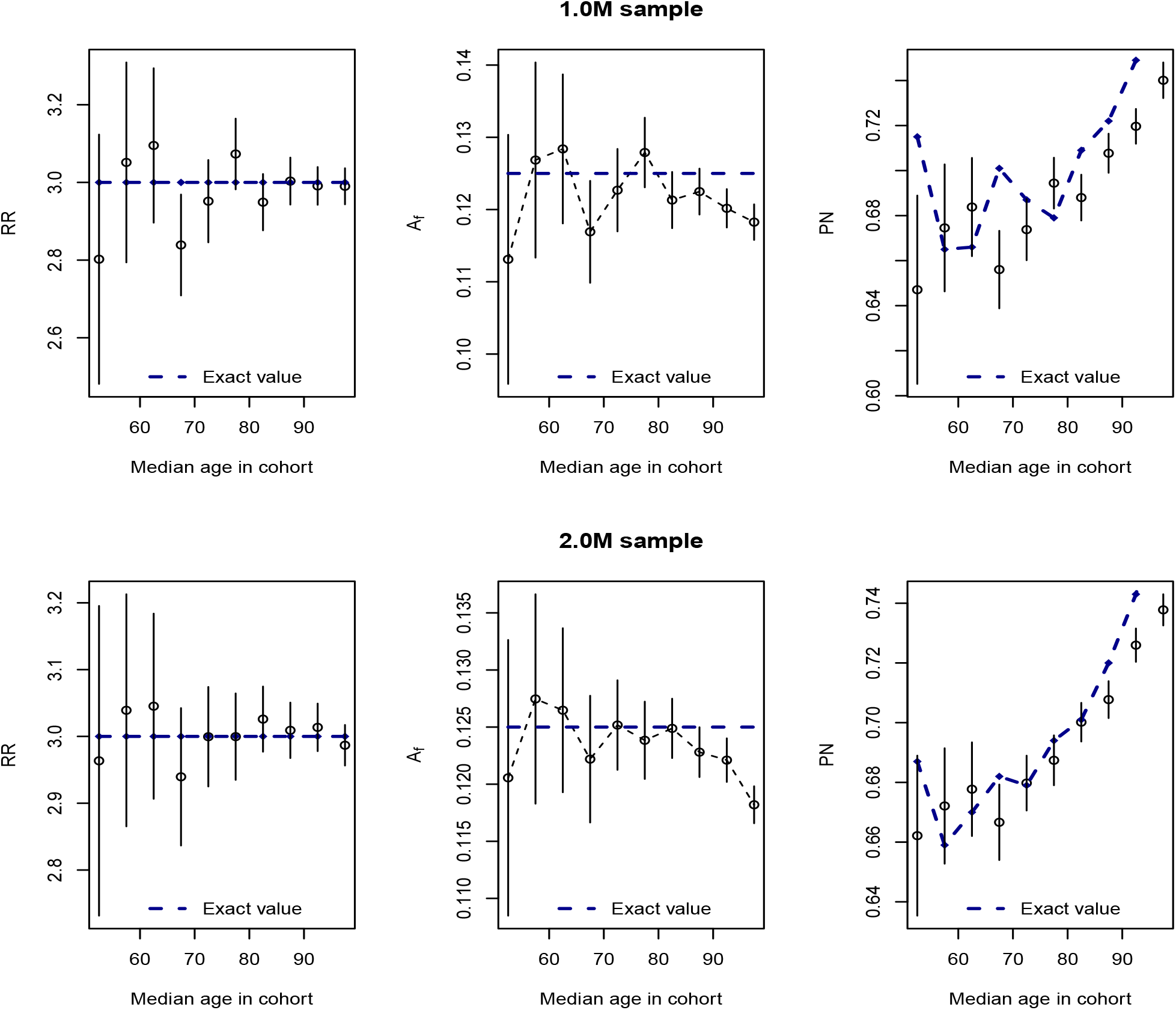
Similar to figure 2, estimates calculated with Eq. 5 are tested with simulated data, here for sample sizes of 1 and 2 million individuals. As in figure 2, the approximation for *A*_*f*_ starts to fail at large enough median cohort age, but the exact value remains within the 95% confidence intervals for ages greater than the median UK life expectancy (∼ 80 years).

Table 1 lists the exact and estimated values for: relative risks RR_0_ and RR, attributable fractions A_*f*0_ and A_*f*_, probability of necessity PN_0_ and PN, the excess risk ratio ERR, and A_*RR*_. The subscripts 0 indicate quantities that are known and calculated exactly, or in the case of PN, expected to be approximated fairly accurately from a combination of data and known values. Although widely discussed, the ERR does not have a clear causal interpretation. For the examples listed in Table 1, ERR has values that are very different to the other causally interpretable quantities. In this example, A_*RR*_ has similar values to PN. A_*f*_ is systematically less than PN, because it is intended to estimate the proportion of disease in a population that in principle could be avoided, and is a product of PN and the probability of exposure in people with the disease (*P* (*X* = 1|*Y* = 1)).

## F Data availability

R Code [18] used to produce the figures and simulated data will be available upon publication. Code used R packages: survival [19], boot [20], meta [21], and xtable [22].

## Notes

### Competing Interest Statement

The authors have declared no competing interest.

### Funding Statement

This study did not receive any funding.

